# Local genetic covariance analysis with lipid traits identifies novel loci for early-onset Alzheimer’s Disease

**DOI:** 10.1101/2024.08.14.24311996

**Authors:** Nicholas R. Ray, Joseph Bradley, Elanur Yilmaz, Caghan Kizil, Jiji T. Kurup, Eden R. Martin, Hans-Ulrich Klein, Brian W. Kunkle, David A. Bennett, Philip L. de Jager, Alzheimer’s Disease Genetics Consortium, Gary W. Beecham, Carlos Cruchaga, Christiane Reitz

**Author notes:** Correspondence: Christiane Reitz, MD PhD Depts. of Neurology, Epidemiology, Sergievsky Center, Taub Institute for Research on the Aging Brain Columbia University 630 W 168th Street New York, NY 10032.

## Abstract

**Background:** The genetic component of early-onset Alzheimer’s disease (EOAD), accounting for ∼10% of all Alzheimer’s disease (AD) cases, is largely unexplained. Recent studies suggest that EOAD may be enriched for variants acting in the lipid pathway.

**Objective:** To examine the shared genetic heritability between EOAD and the lipid pathway by genome-wide multi-trait genetic covariance analyses.

**Methods:** Summary statistics were obtained from the GWAS meta-analyses of EOAD by the Alzheimer’s Disease Genetics Consortium (*n*=19,668) and five blood lipid traits by the Global Lipids Genetics Consortium (*n*=1,320,016), and genetic covariance analyses were performed via SUPERGNOVA. Genes in linkage disequilibrium (LD) with top EOAD hits in identified regions of covariance with lipid traits were scored and ranked for causality by combining evidence from gene-based analysis, AD-risk scores incorporating transcriptomic and proteomic evidence, eQTL data, eQTL colocalization analyses, DNA methylation data, and single-cell RNA sequencing analyses.

**Results:** Local genetic covariance analyses identified 3 regions of covariance between EOAD and at least one lipid trait. Gene prioritization nominated 3 likely causative genes at these loci: *ANKDD1B*, *CUZD1*, and *MS4A64*.

**Conclusion:** The current study identified genetic covariance between EOAD and lipids, providing further evidence of shared genetic architecture and mechanistic pathways between the two traits.

## INTRODUCTION

Alzheimer’s Disease (AD) is a highly prevalent progressive neurodegenerative disorder that places a substantial physical and emotional burden on patients and caregivers, and a significant financial toll on health care and social care systems(1). While most AD patients are elderly individuals, 5-10% of cases occur before the age of 65 years and are classified as early-onset Alzheimer’s Disease (EOAD)(2). EOAD has a substantial genetic basis with a heritability of 91% to 100%(3). Studies of multiplex families with EOAD led to the identification of AD-causing mutations in the amyloid precursor protein (*APP*), presenilin 1 (*PSEN1*), and presenilin 2 (*PSEN2*) genes, playing a pivotal role in the implementation of the amyloid hypothesis in AD, which proposes an increase in β-amyloid production as a causative mechanism in AD etiology(4). However, while the exact contribution of variation in *APP*, *PSEN1*, and *PSEN2* to EOAD prevalence is unknown, it is estimated to be less than 10% of all incident EOAD cases, leaving ∼90% of EOAD cases unexplained(3–5). A large proportion of EOAD heritability is expected to be explained by SNPs that do not pass the Bonferroni-corrected significance threshold(6).

Identification of the remaining genetic variation underlying EOAD and mapping of the mechanistic pathways involved is critical to disentangle the substantial clinical heterogeneity observed in this trait, develop prediction models, and develop more effective targets for screening, prevention, and treatment. A powerful approach to identify additional causative variants and biological pathways underlying complex traits are multi-trait analyses estimating local genetic covariance (i.e. genetic similarity in specific genomic regions) with other traits potentially sharing etiologic mechanisms. Acknowledging the importance of disentangling pleiotropy to pinpoint disease etiology and potentially reposition drugs for complex diseases, multi-trait modeling has recently undergone rapid developments and has resulted in significantly improved methods. To identify new genetic loci underlying EOAD, we examined genome-wide local genetic covariance with five lipid traits: total cholesterol (TC), high-density lipoprotein cholesterol (HDL-C), low-density lipoprotein cholesterol (LDL-C), non-high-density lipoprotein cholesterol (nonHDL-C), and triglycerides (TG). A large body of epidemiologic studies of AD by us(7, 8) and others(9–15) has shown that cholesterol levels elevated in midlife increase the risk of AD and cognitive decline, and associations of higher LDL-C with increased cerebral β-amyloid load have been observed in autopsy and in vivo imaging studies(16, 17). Similarly, AD risk is lower among statin users, and this association appears to be more pronounced with longer treatment exposure and the use of more potent drugs(18–23), although corresponding observational data on other lipid-lowering drug classes are limited and ambiguous(24). Studies specifically examining the association with EOAD are scarce, but a recent study of plasma samples from 2,125 EOAD cases and controls observed an association between EOAD and higher levels of LDL-C independent of the effects of *APOE*; as well as an enrichment of rare coding variants of *APOB*, a gene known to influence plasma cholesterol levels(25). This supports the notion that EOAD may share common genetic loci with the lipid pathway. Following genetic covariance analyses, we validated putative loci observed to be shared between EOAD and any of the five lipid traits by conducting gene-based analysis; extracting publicly available AD risk scores calculated from genome-wide association studies (GWAS) and expression quantitative trait locus (eQTL), transcriptomic, and proteomic data; examining publicly available data from brain eQTL studies; performing colocalization analyses between EOAD summary statistics and eQTL data; inspecting brain DNA methylation data from the Religious Orders Study and Rush Memory and Aging Project (ROSMAP) cohorts; and analyzing single cell RNA sequencing data from both humans and zebrafish.

## METHODS

### EOAD and lipid trait GWAS studies

Quality-controlled, ancestry-specific summary statistics were obtained from the GWAS meta-analyses of EOAD by the Alzheimer’s Disease Genetics Consortium (*n* = 19,668) and plasma lipid levels conducted by the Global Lipids Genetics Consortium (*n* = 1,320,016).

#### EOAD GWAS

In brief, the EOAD GWAS included genetic data on 6,282 EOAD cases with AD diagnosis at or before age 70 and 13,386 cognitively normal controls with age at examination greater than 70(26). Participants were obtained from 47 independent datasets assembled through the Alzheimer’s Disease Genetics Consortium (ADGC), and European ancestry was determined by genetic principal component analysis. Demographic information for each cohort is described in Supplementary Table 1. Genetic data was genotyped on multiple genotyping arrays, QCed, imputed using the TOPMed imputation server, and aligned to the GRCh38 reference panel(26).

#### Lipids GWAS

Data on the genetic architecture of lipid traits were derived from Graham et al. (2021) which meta-analyzed HDL-C, LDL-C, non-HDL-C, TC and TG summary statistics from 1,654,960 individuals across 201 individual studies(27). Data from each cohort was QCed and imputed to the 1000 Genomes Phase 3 v5 (1KGP3) and the Haplotype Reference Consortium (HRC) panel(27, 28). The genetic covariance analyses presented here were conducted on ancestry-specific summary statistics obtained from 1,320,016 individuals of European ancestry.

### Analysis of genetic covariance between EOAD and lipid traits

Estimation of genetic covariance of EOAD with individual lipid traits (TC, HDL-C, nonHDL-C, LDL-C, and TG) was performed via SUPERGNOVA(29). SUPERGNOVA estimates local genetic correlation while taking into account linkage disequilibrium structure by decorrelating local z-scores with eigenvectors of the local LD matrix followed by estimation of local genetic covariance through a weighted least squares regression in each region. This technique has been demonstrated to be superior to other available methods such as LD score regression (LDSC) or GeNetic cOVariance Analyzer (GNOVA)(29). The genome was partitioned into 2,353 approximately independent regions (∼1.6 centimorgan on average) using LDetect(30), and LD was estimated using the 1000 Genomes Phase3 European reference panel(31). Accordingly, the *P*-value threshold for significance of local genetic covariance between EOAD and each lipid trait was set based on a conservative Bonferroni threshold of *P* < 2.12×10^-5^ (0.05/2,353 bins). Regions resulting from the genetic covariance analyses were only reported if a strong genetic signal was observed in both traits, as defined by a *P*-value cutoff of 5×10^-5^. In addition, within-trait local genetic association and haplotype structure at these loci was assessed in the respective individual trait summary statistics (EOAD and the respective lipid trait showing genetic covariance), and visualized, annotated, and aligned across traits using LocusZoom(32).

### Gene prioritization

Genes included for prioritization were selected from the regions showing genetic correlation between EOAD and any of the five lipid traits. Locus zoom plots of these regions were inspected visually and any gene within LD of the EOAD top SNP in each region (any part of the gene can be within the LD block) was investigated further and given a composite score based on: 1) results from gene-based analysis; 2) AD risk scores from *AGORA* (https://agora.adknowledgeportal.org/about) hosting high-dimensional human transcriptomic, proteomic, and metabolomic evidence for gene association with AD; 3) MetaBrain eQTL data; 4) eQTL colocalization analyses; 5) ROSMAP brain DNA methylation data (see below); and 6) single cell RNA sequencing data from both humans and zebrafish. The resulting composite scores were used to nominate the most likely causative gene(s) in each region by selecting the top scoring gene(s) from each region.

#### Gene-based analysis

We performed gene-based analysis on our EOAD summary statistics using the MAGMA(33) software via the FUMA(34) web tool (https://fuma.ctglab.nl/). The 1000 Genomes Phase3 European reference panel(31) was employed along with the following parameters: FUMA v1.6.0; MAGMA v1.08; *P*-value of lead SNPs < 1×10^-5^; *P*-value of GWAS SNPs < .05; r^2^ threshold to define independent significant SNPs ≥ 0.6; second r^2^ threshold to define lead SNPs ≥ 0.1; minimum MAF = 0; maximum distance between LD blocks = 250kb. A window was set 35kb upstream and 10kb window downstream of the gene. 19163 genes were tested by MAGMA, resulting in a Bonferroni-corrected *P*-value threshold of 2.61×10^-6^.

#### AD risk score data

Genetic, multi-omic, and target AD risk scores were extracted for each gene of interest from the Agora tool (https://agora.adknowledgeportal.org/). The *Genetics Risk Score* ranges from 0-3 and is based on: 1) data from 24 GWAS or GWAS by proxy studies and 3 QTL studies; 2) phenotypic evidence supporting a gene from human and/or animal models; and 3) whether a gene has a model in development through the MODEL-AD consortium (https://www.model-ad.org/)(35). The *Multi-omic Risk Score* ranges from 0-2 and is based on: 1) transcriptomic data from RNA-Seq profiling from 8 neocortical tissues and 2) proteomic data from label-free quantitation (LFQ) and Tandem Mass Tagging (TMT) shot-gun profiling methods generated from 8 neocortical tissues(35). The *Target Risk Score* ranges from 0-5 and is a sum of the Genetic Risk Score and the Multi-omic Risk Score.

#### MetaBrain cis-eQTL data

Results from *cis*-eQTL analyses from the cortex (n = 2,683) and hippocampus (n = 208) of individuals of European ancestry were obtained from MetaBrain (www.metabrain.nl). MetaBrain has collected 6,518 genotype samples and 8,613 bulk RNA-sequenced samples across 14 datasets and has performed ancestry and brain region specific *cis*- and *trans*-eQTL metanalyses(36), defining cis-eQTLs as common variants (MAF >1%) within 1 megabase (Mb) of the transcription start site of a protein-coding gene. For the present study, *cis*-eQTLs were extracted from this data for each gene under the peaks of the 3 regions resulting from the genetic covariance analyses, and the eQTL with the minimum *P*-value was selected to represent each gene.

#### eQTL colocalization analyses

To calculate the probability that genes in identified regions of covariance between EOAD and at least one lipid trait share a single causal variant with eQTL loci (this probability is referred to as H4), we performed colocalization analyses between the regions resulting from our genetic covariance analyses and a set of 61 eQTL datasets (Supplementary Table 2) using the ‘coloc’ package in R(37–39).

#### Brain DNA methylation analyses in the ROS/MAP cohort

DNA methylation data came from frozen dorsolateral prefrontal cortex from 761 participants in ROSMAP(40). All ROSMAP participants enroll without known dementia, agree to annual clinical evaluation and agree to brain donation at the time of death. Both studies were approved by an Institutional Review Board of Rush University Medical Center. All participants signed informed and repository consents and an Anatomic Gift Act. Details of the data generation have been previously published in detail whereby methylation profiles were generated using the Illumina HumanMethylation450 beadset(41, 42). For the present analyses the β-values reported by the Illumina platform for each probe ranging from 0 (no methylation) to 1 (100% methylation) were utilized as the methylation level measurement for the targeted CG site in a given sample. To examine identified top loci for differentially methylated regions associated with AD pathology, we used a linear model, adjusting for age at death, sex, experimental batch, and bisulfite conversion efficiency. β-amyloid load and PHF-tau tangle density were generated as previously described(43).

#### Single cell RNA sequencing

To evaluate the RNA expressions of target genes at the single nucleus/cell level, we evaluated publicly available human single cell sequencing data (GSE157827)(44) and analyzed zebrafish data generated in-house. For human snRNA, we selected the single cell expression matrices of 4 AD and 4 control samples that were matched on sex and age. Matrices were generated with 10X function of the ‘Seurat’ (version 4.1.3) R package(45). To create the Seurat object, we filtered out any cells with less than 200 expressed genes, and with genes expressed in less than 3 cells. Following the normalization of the dataset, the top 2,000 genes were used for further analyses. Anchors were identified with the *FindIntegrationAnchors* function and integration was performed using the *IntegrateData* function. We used ‘DoubletFinder’(46) to remove doublets and performed the rest of the analyses on singlets only. The integrated Seurat object included 44,132 cells (27,198 for AD, and 16,934 for Control) with 29,772 features. The data were scaled using all genes, and 30 PCAs (RunPCA) were identified using the *RunPCA* function in the ‘Seurat’ package(45). Thirty clusters were identified with resolution 1. The main cell types were defined using *AQP4* and *GFAP* for astroglia; *SLC17A7* and *NRGN* for excitatory neurons; *GAD1* and *GAD2* for inhibitory neurons; *PDGFRB*, *MCAM* and *GRM8* for pericytes; *C3* and *DOCK8* for microglia; *PLP1* and *MOBP* for oligodendrocytes; *PDGFRA* and *VCAN* for OPC; and *FLIT1* and *CLDN5* for endothelial cells.

We used the same methods and parameters as described above for creating a Seurat object with our zebrafish dataset. The main cell types were identified as described elsewhere(47, 48). Briefly, we used *s100b* and *gfap* for astroglia; *sv2a*, *nrgna*, *grin1a*, *grin1b* for neurons; *pdgfrb* and *kcne4* for pericytes; *cd74a* and *apoc1* for microglia; *mbpa* and *mpz* for oligodendrocytes; *aplnra* for OPC; *myh11a* and *tagln2* for vascular smooth muscle cells; *lyve1b* for lymph endothelial cells; and *kdrl* for vascular cells. To find the average expression of the genes, we used *AverageExpression* function. To define the percent expression of the given genes, *PrctCellExpringGene* was used.

Differential gene expression (DEG) in clusters was performed using the *FindMarkers* function by comparing AD cases against age-matched controls in humans and zebrafish injected with Aβ42 (AD model) versus those injected with phosphate buffered saline (control group). *P*-values and log2 fold change values of DEG results were transformed to generate the DEG index using the following equation: -log10(*P*) × (1 - |log2 fold change value|). If a gene is differentially expressed in multiple cell clusters, the one with lowest *P*-value was selected for the index. If no DEG was observed for a gene, a non-DEG penalty value of .25 was assigned for that gene.

#### Composite scores for each gene

*P*-values were extracted and transformed – using -log10(*P*) – from the results of the MetaBrain eQTL data, the gene-based analysis, and the ROSMAP methylation data for each gene. Where results existed for multiple variants within a gene, the variant with the minimum *P*-value was selected to represent the gene. The target risk score for each gene was extracted from Agora. After performing the colocalization analyses between the EOAD summary statistics and the multiple eQTL datasets, the maximum H4 value between EOAD and the various eQTL datasets was selected to represent each gene of interest. A score was calculated for each gene for both the human and zebrafish single-cell RNA sequencing analyses by multiplying the average proportion of each gene’s expression across different brain cell types by the average amount of each gene’s expression across the same cell types and by each gene’s DEG index. Z-scores were calculated for each of the variables described above, subsequently these z-scores were summed to form a total prioritization score for each gene. Any genes with scores in the top 10^th^ percentile for each region resulting from the genetic covariance analyses were nominated as likely causative genes for that locus.

## RESULTS

### Analysis of genetic covariance between EOAD and lipid traits

Top results of the genetic covariance analyses with the five lipid traits (TC, HDL-C, LDL-C, nonHDL-C, and TG) are summarized in Table 1, with corresponding regional association plots displayed in Supplementary Figures 1-3. All base-pair positions in this section refer to build GRCh37. Genetic covariance analyses of EOAD with each of the five lipid traits identified 3 regions showing genetic correlation between EOAD and at least one of the five lipid traits at *P* < 2.12×10^-5^. The region showing strongest genetic correlation is located on chromosome 5q13.3 at 73,508,509-75,240,469 bp and showed genetic covariance between EOAD and TC (*P* = 5.5×10^-10^), LDL-C (*P* = 8.17×10^-9^), and nonHDL-C (*P* = 1.30×10^-10^) (Table 1; Supplementary Figure 1). A second region on chromosome 10q26.13 at 123,855,124-124,894,743 bp showed genetic covariance between EOAD and TC (*P* = 3.78×10^-9^), LDL-C (*P* = 1.26×10^-8^), and nonHDL-C (*P* = 2.90×10^-9^) (Table 1; Supplementary Figure 2). And lastly, a region on chromosome 11q12.1-q12.2 at 59,620,206-61,870,732 bp showed genetic covariance between EOAD and nonHDL-C (*P* = 1.60×10^-6^) (Table 1; Supplementary Figure 3). Examination of LD patterns in these regions identified in total 26 genes under the peaks. A complete list of these genes in each region can be found in Supplementary Table 3.

**Table 1.**
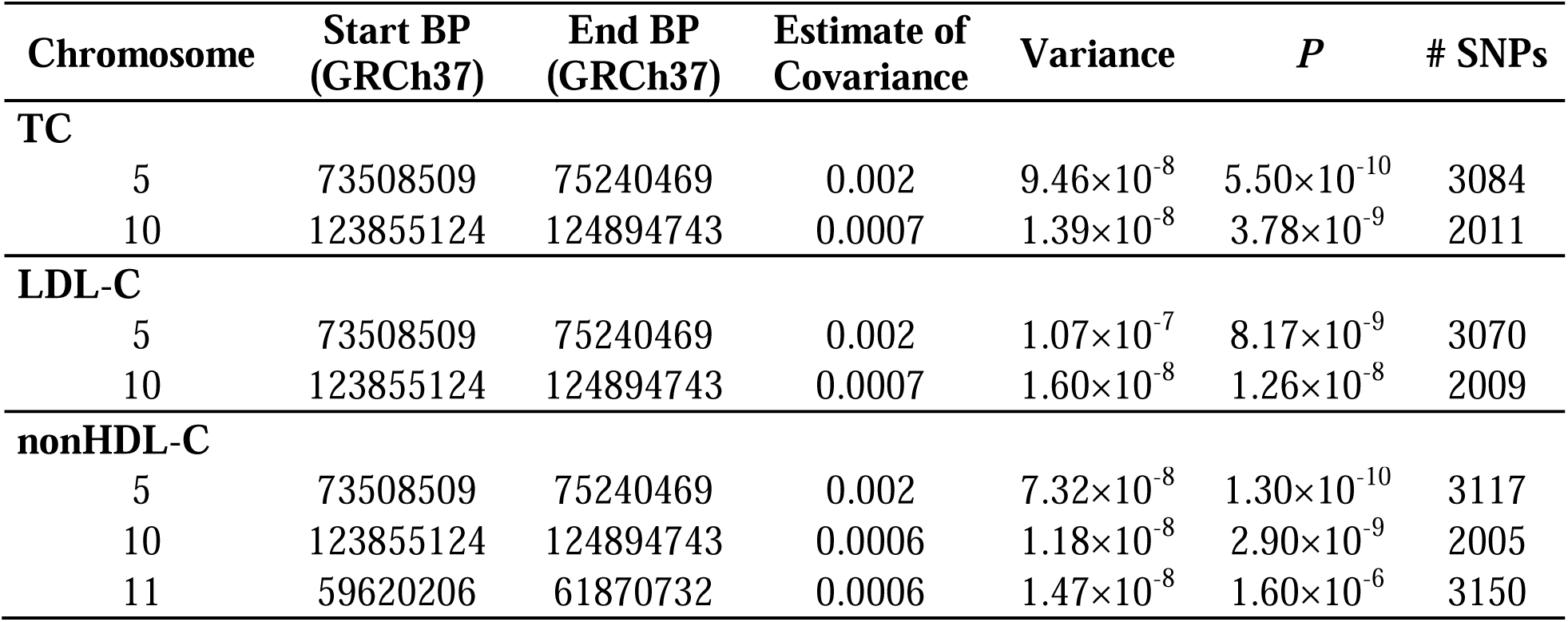
Results of genetic covariance analyses for regions with *P* < 2.12×10^-5^.

| Chromosome | Start BP<br>(GRCh37) | End BP<br>(GRCh37) | Estimate of<br>Covariance | Variance | $P$ | # SNPs |
| --- | --- | --- | --- | --- | --- | --- |
| **TC** |  |  |  |  |  |  |
| 5 | 73508509 | 75240469 | 0.002 | $9.46 \times 10^{-8}$ | $5.50 \times 10^{-10}$ | 3084 |
| 10 | 123855124 | 124894743 | 0.0007 | $1.39 \times 10^{-8}$ | $3.78 \times 10^{-9}$ | 2011 |
| **LDL-C** |  |  |  |  |  |  |
| 5 | 73508509 | 75240469 | 0.002 | $1.07 \times 10^{-7}$ | $8.17 \times 10^{-9}$ | 3070 |
| 10 | 123855124 | 124894743 | 0.0007 | $1.60 \times 10^{-8}$ | $1.26 \times 10^{-8}$ | 2009 |
| **nonHDL-C** |  |  |  |  |  |  |
| 5 | 73508509 | 75240469 | 0.002 | $7.32 \times 10^{-8}$ | $1.30 \times 10^{-10}$ | 3117 |
| 10 | 123855124 | 124894743 | 0.0006 | $1.18 \times 10^{-8}$ | $2.90 \times 10^{-9}$ | 2005 |
| 11 | 59620206 | 61870732 | 0.0006 | $1.47 \times 10^{-8}$ | $1.60 \times 10^{-6}$ | 3150 |

### Gene prioritization at identified loci showing genetic covariance

To identify likely causative genes in each of the 3 regions showing genetic covariance between EOAD and lipid traits, we calculated composite scores for each gene from gene-based analysis, Agora AD-risk scores, MetaBrain eQTL data, eQTL colocalization analyses, ROSMAP brain methylation data, and single cell RNA sequencing analyses in humans and zebrafish. Results from the gene-based analysis are displayed in Supplementary Table 4. Target Risk Scores extracted from Agora (https://agora.adknowledgeportal.org/) for each gene of interest are shown in Supplementary Table 5. Results from the MetaBrain cis-eQTL mapping for the 26 genes of interest with the minimum *P*-value for each gene are detailed in Supplementary Tables 6 and 7 respectively. Max H4 values from the coloc eQTL colocalization analyses were extracted for each gene of interest along with their associated eQTL datasets (Supplementary Table 8). The variant with the minimum methylation *P*-value for each gene was extracted separately for both β-amyloid load and tau tangle density, and the results of the methylation analyses for these variants are shown in Supplementary Tables 9 and 10, respectively. Results of the single-cell RNA sequencing analyses for humans and zebrafish, where a total score for each gene was derived by multiplying average proportion of expression by average level of expression by DEG index, are detailed in Supplementary Tables 11 and 12, respectively.

Z-scores were calculated and summed for the variables mentioned above to create a composite “causality” score for each gene (Supplementary Table 13). Boxplots for these resulting scores for each significant region from the genetic covariance analyses are shown in Figure 1. Three genes were nominated as likely to be causal across the 3 regions: *ANKDD1B*, *CUZD1*, and *MS46A*. *ANKDD1B* is the highest scoring gene in the region on chromosome 5q13.3 and is nominally associated with EOAD according to gene-based analysis (*P* = 2.9×10^-4^; Supplementary Table 4). Meta-analyses of *cis*-eQTLs for brain-related traits show at least one variant in the *ANKDD1B* gene to be highly significant in the cortex (*P* = 3.18×10^-110^; Supplementary Table 6) and nominally significant in the hippocampus (*P* = .003; Supplementary Table 7)(36). *ANKDD1B* has an AD target risk score of 1.62 according to Agora (https://agora.adknowledgeportal.org/; Supplementary Table 5), and colocalization analyses report a high probability that EOAD and eQTL data share a causal variant in the *ANKDD1B* gene (H4 = .82; Supplementary Table 8).

**Figure 1.**
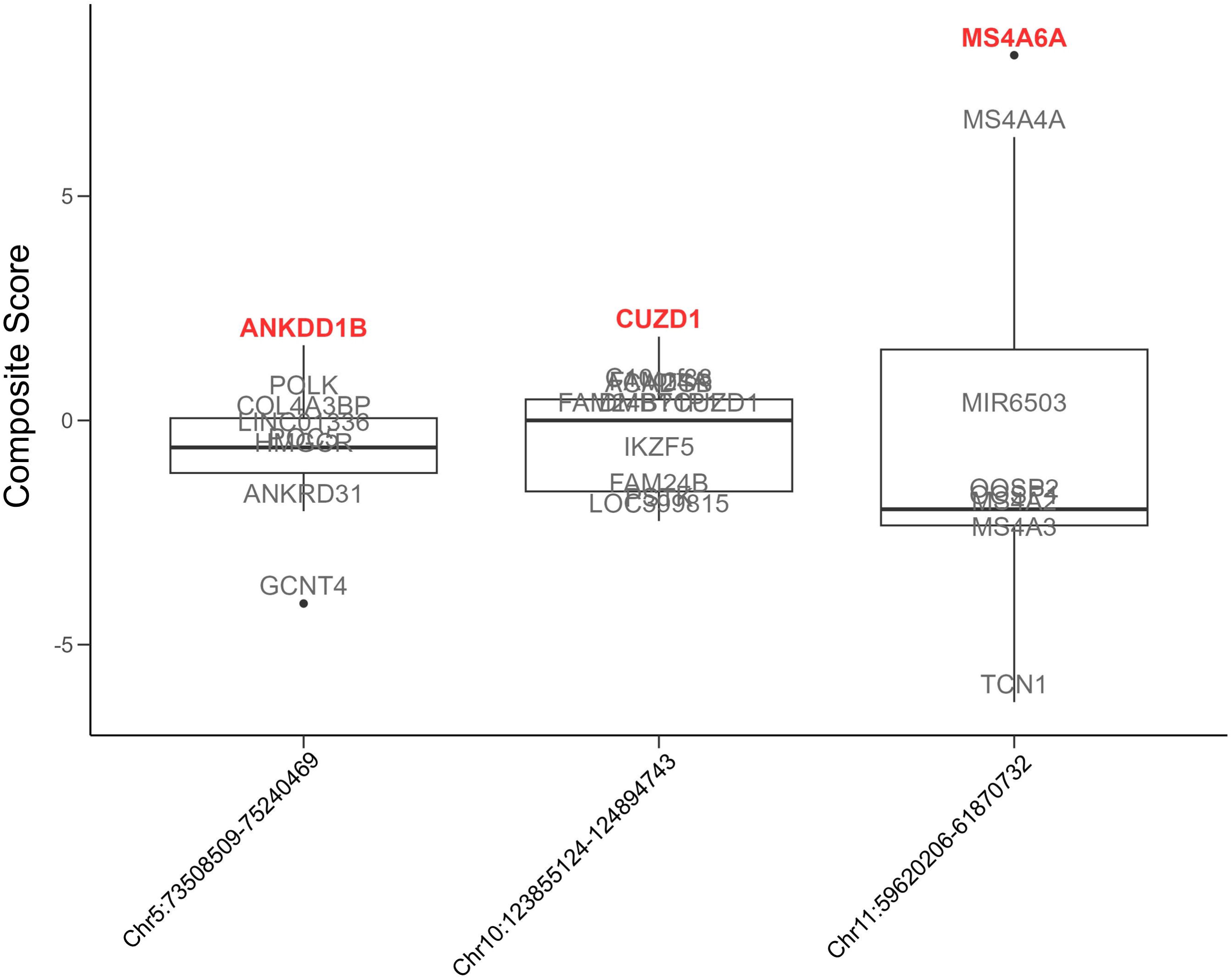
Boxplot of composite scores for all genes in each region resulting from the genetic covariance analyses between EOAD and lipid traits. The chromosome and base-pair start and end positions for each region are displayed on the x-axis using genome build GRCh37. Genes with composite score values in the top 10^th^ percentile for each region are bolded and shown in red, while the rest of the genes are shown in grey.

The highest scoring gene in the region on chromosome 10q26.13 is *CUZD1*. Gene-based analysis shows *CUZD1* to be nominally associated with EOAD (*P* = .01; Supplementary Table 4) and at least one variant in the *CUZD1* gene is a *cis*-eQTL in the cortex (*P* = 1.05×10^-91^; Supplementary Table 6) and the hippocampus (*P* = 1.94×10^-7^; Supplementary Table 7)(36). Colocalization analyses report a high probability that EOAD and eQTL data share a causal variant in the *CUZD1* gene (H4 = .84; Supplementary Table 8) and Agora scores the AD risk of the gene at 1.48 (https://agora.adknowledgeportal.org/; Supplementary Table 5).

Finally, the highest scoring gene in the region on chromosome 11q12.1-q12.2 is *MS46A*, which is highly significant according to gene-based analysis (*P* = 4.07×10^-10^; Supplementary Table 4) and has a high AD risk score of 3.49 (https://agora.adknowledgeportal.org/; Supplementary Table 5). At least one variant in *MS46A* is a significant eQTL in the cortex (*P* = 4.26×10^-6^; Supplementary Table 6) and the hippocampus (*P* = 6.15×10^-4^; Supplementary Table 7)(36), and EOAD is likely to share a causal variant with eQTLs in the gene according to colocalization analysis (H4 = .96; Supplementary Table 8). Analysis of brain methylation data report that at least one variant in *MS46A* is nominally significant for both amyloid (*P* = .02; Supplementary Table 9) and tau (*P* = .005; Supplementary Table 10) methylation; and single cell RNA sequencing analysis shows that *MS46A* is differentially expressed between AD cases and controls in humans (*P* = 3.74×10^-20^; Supplementary Table 11).

## DISCUSSION

To identify genetic regions and mechanistic pathways that are shared between early-onset EOAD and dyslipidemia, we conducted hypothesis-free genetic covariance analyses between EOAD and lipid traits. These analyses identified 3 genetically shared regions on chromosomes 5q13.3, 10q26.13, and 11q12.1-q12.2. Construction of composite scores integrating gene-based analyses and a wealth of multi-omics data prioritized 3 genes in these regions most likely to be causal: *ANKDD1B*, *CUZD1*, and *MS46A*. All these identified genes act in mechanistic pathways related to AD and/or have been associated with AD related read-outs in animal, cell biological, or neuropathological studies.

*ANKDD1B*, the highest scoring gene in the region on chromosome chr5q13.3, encodes the ankyrin repeat and death domain containing 1 B protein. *ANKDD1B* has been shown to be associated with dyslipidemia (specifically LDL-C)(49) and type 2 diabetes(50) in major GWAS studies, and is one of 28 genes included in a recent polygenic risk score for severe hypercholesterolemia (defined as LDL-C > 4.9 mmol/L)(51). *ANKDD1B* was reported as one of two top genes connecting migraines and major depressive disorder in a genetic correlation analysis(52) and is hypermethylated in response to low-dose lead exposure in mice(53). Diabetes, depression, and lead exposure have all been linked with cognitive decline and various neurological disorders including AD(54–59).

*CUZD1* is the highest scoring gene in the region on chromosome 10q26.13 and encodes a protein located in secretory granules in the pancreas that is thought to affect lipid-related metabolite levels(60). *CUZD1* is a contributing gene to the zymogen activation pathway, which is enriched in the top 5% of genes associated with AD from a genome-wide meta-analysis(61). Increased levels of the CUZD1 protein have also been correlated with genetic risk of migraine(62).

The highest scoring gene in the region on chromosome chr11q12.1-q12.2 is *MS4A6A*, a known AD risk gene that encodes a member of the membrane-spanning 4A gene family(63). Several meta-analyses have found that a variant (rs610932) in *MS4A6A* correlates with decreased risk for AD(63–65). Colocalization analysis identified a shared causal variant in *MS4A6A* affecting a locus near *MS4A4A* in one of the most recent AD GWAS(66). A transcriptome-wide association study (TWAS) found that increased expression of *MS4A6A* in monocytes associated with AD risk(67) and a DNA methylation study suggested that *MS4A6A* expression may mediate AD risk(68). *MS4A6A* is also involved in the formation of atherosclerosis(69) and is differentially associated with various classes of lipids between AD cases and controls(70).

This study has several strengths. To our knowledge this is the first study assessing genetic covariance between the early-onset form of AD and the lipid pathway. AD cases and controls in the parent EOAD GWAS were derived from datasets with meticulous characterization for cognitive impairment, age at onset and AD. In addition, to further validate shared identified loci, this study was able to capitalize on a variety of omics data from various independent sources, providing significant supportive evidence for the plausibility of candidate genes at identified loci. The evidence presented here is based on correlational analyses, however, and functional follow-up for these genes is required to determine causality. In addition, it is possible that regions of genetic covariance were missed in our study due to a lack of statistical power (particularly in the EOAD dataset) or lack of data from sex-stratified analyses.

In summary, this study suggests that EOAD shares genetic heritability with the lipids pathway, and that the common genetic loci include the *ANKDD1B, CUZD1,* and *MS4A6A* genes. These genes could lead to improved screening, prevention, and treatment for AD by targeting shared mechanistic pathways between EOAD and lipids. Future studies are needed that clarify the molecular mechanisms through which these genes modulate risk of EOAD, and that identify the specific causative variants at these and additional remaining loci that underlie the contribution of lipid metabolism to AD pathogenesis.

## Data Availability

The EOAD summary statistics will be publicly available upon publication of the relevant paper here: https://adsp.niagads.org/
The lipids summary statistics are available here: https://csg.sph.umich.edu/willer/public/glgc-lipids2021/

## ACKNOWLEDGEMENTS

This study was supported by NIH grants R01AG064614 (CR, GB, CC), RF1AG054080 (CR, GB), P30AG066462 (CR), R01AG044546 (CC), P01AG003991(CC), RF1AG053303 (CC), RF1AG058501 (CC), U01AG058922 (CC)), U01AG032984 (GS, LSW), U54AG052427 (GS, LSW), the Alzheimer Association (NIRG-11-200110 (CC), BAND-14-338165 (CC), AARG-16-441560 (CC) and BFG-15-362540 (CC).

ROSMAP is supported by P30AG10161, P30AG72975, R01AG15819, R01AG17917, U01AG46152, and U01AG61356. ROSMAP resources can be requested at https://www.radc.rush.edu and www.synpase.org.

## Competing Interests Statement

CC receives research support from: Biogen, EISAI, Alector and Parabon. The funders of the study had no role in the collection, analysis, or interpretation of data; in the writing of the report; or in the decision to submit the paper for publication. CC is a member of the advisory board of Vivid genetics, Halia Therapeutics and ADx Healthcare.

## Notes

### Competing Interest Statement

Carlos Cruchaga receives research support from: Biogen, EISAI, Alector and Parabon. The funders of the study had no role in the collection, analysis, or interpretation of data; in the writing of the report; or in the decision to submit the paper for publication. Carlos Cruchaga is a member of the advisory board of Vivid genetics, Halia Therapeutics and ADx Healthcare.

### Author Declarations

The EOAD summary statistics will be publicly available upon publication of the relevant paper here: https://adsp.niagads.org/ The lipids summary statistics are available here: https://csg.sph.umich.edu/willer/public/glgc-lipids2021/

